# Assessing feasibility and risk to translate, de-identify and summarize medical reports using deep learning

**DOI:** 10.1101/2023.07.27.23293234

**Authors:** Lucas W. Gauthier, Marjolaine Willems, Nicolas Chatron, Camille Cenni, Pierre Meyer, Valentin Ruault, Constance Wells, Quentin Sabbagh, David Genevieve, Kevin Yauy

## Abstract

**Background:** Precision medicine requires accurate phenotyping and data sharing, particularly for rare diseases. However, sharing medical reports across language barriers is challenging. Alternatively, inconsistent and incomplete clinical summary provided by physicians using Human Phenotype Ontology (HPO) can lead to a loss of clinical information.

**Methods:** To assess feasibility and risk of using deep learning methods to translate, de-identify and summarize medical reports, we developed an open-source deep learning multi-language software in line with health data privacy. We conducted a non-inferiority clinical trial using deep learning methods to de-identify protected health information (PHI) targeting a minimum sensitivity of 90% and specificity of 75%, and summarize non-English medical reports in HPO format, aiming a sensitivity of 75% and specificity of 90%.

**Results:** From March to April 2023, we evaluated 50 non-English medical reports from 8 physicians and 12 different groups of diseases, which included neurodevelopmental disorders, congenital disorders, fetal pathology and oncology. Reports contain in median 15 PHI and 7 HPO terms. Deep learning method achieved a sensitivity of 99% and a specificity of 87% in de-identification, and a sensitivity of 78% and a specificity of 92% in summarizing medical reports, reporting an average number of 6.6 HPO terms per report, which is equivalent to the number of HPO terms provided usually by physicians in databases (6.8 in PhenoDB).

**Conclusions:** De-identification and summarization of non-English medical reports using deep learning methods reports non-inferior performance, providing insights on AI usage to facilitate precision medicine.

**Graphical abstract:** Illustration of the non-inferiority trial for de-identification and summarization of non-english medical reports and main statistical performances.

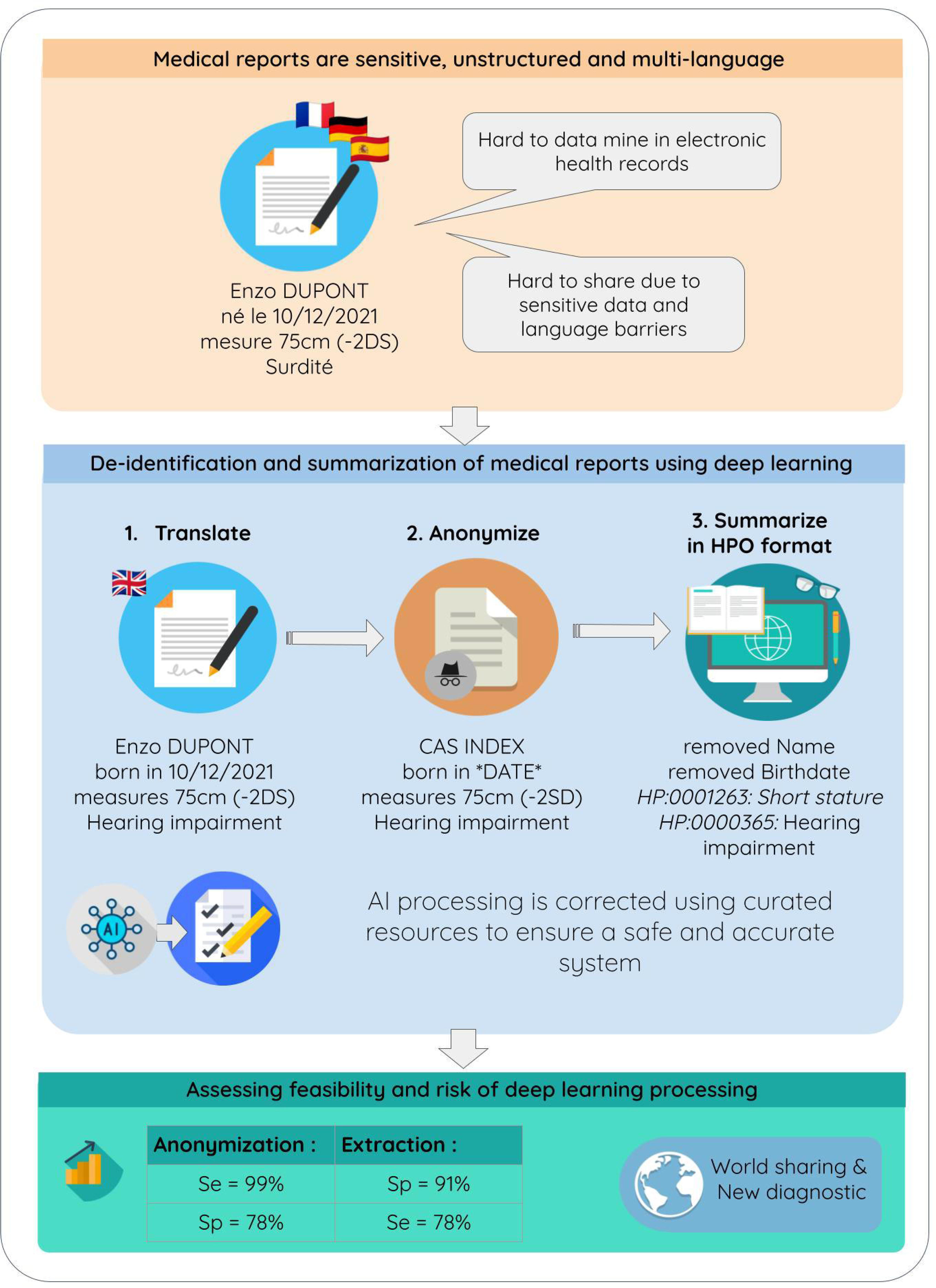

## Introduction

Precision medicine requires precise phenotyping and structured electronic health records based on clinical data sharing, especially in the context of rare diseases, where matching patients worldwide is crucial ^1^.

In particular, medical reports contain critical information about a patient’s condition, making them crucial for communication between healthcare providers ^2^. However, sharing reports between providers who speak different languages can be challenging and time-consuming, especially if the reports need to be translated and de-identified to protect patient privacy ^3^. Morever, medical reports are unstructured text which is difficult to exploit in precision medicine.

As an alternative, the community adopted the Human Phenotype Ontology (HPO) that enabled physicians to use a common language with machines ^4^. Sharing HPO terms summarizing clinical descriptions has already proved effective in discovering new diseases through MatchMaker Exchange^5^ and is a key element for computational phenotype analysis in genome sequencing analysis ^6^. However, HPO terms furnished by physicians appear to have significant differences when evaluated, requiring standardization of practices and reproducibility to communicate more efficiently. Moreover, they are generally partially filled, losing clinical information ^7^.

Recent advancements in Artificial Intelligence (AI) including deep learning and natural language processing show potential in addressing sharing of medical information challenges ^8^. However, the use of black box approach and cloud-only systems may not comply with the General Data Protection Regulation (GDPR) with health data. Therefore, it is recommended to keep Protected Health Informations (PHI) in-house and keep processing of medical data off-line ^9^. Achieving de-identified and structured clinical data is a key element to efficiently exploit clinical data warehouse and apply algorithms to discover and better understand diseases. To assess the feasability and the risk to translate, de-identify and summarize medical reports using deep learning, we developed an open-source software for multi-language translation, de-identification, and summarization of medical reports using HPO terms in line with FAIR principles while ensuring data privacy and security ^10^. This deep learning-based software was evaluated by a non-inferiority trial of 50 medical reports in de-identification and summarization using HPO terms compared to a physician.

## Methods

### STUDY POPULATION

A multi-centric and prospective study was conducted at University Hospital Center of Montpellier and University Hospital Center of Nîmes to evaluate the performance of a software that generates English translations, de-identifies, and summarizes symptoms in the Human Phenotype Ontology (HPO) format from non-english medical consultation or hospitalization reports. The study included patients with suspected genetic disorders and non-opposition consent for the use of their clinical data in this study was collected after being informed by physicians. The first edited or available reports (consultation or hospitalization) in patient’s Electronic Health Records (EHR) were included and their first and last name were pseudonymised before inclusion. Patients who did not provide consent or did not have consultation or hospitalization report were excluded. The study was approved by the Institutional Review Board (IRB) of University Hospital Center of Montpellier on April 14th 2023 (IRB-MTP_2023_04_202301351).

### STUDY DESIGN

In this study, a physician examined 50 medical reports and initiated a multi-step analysis using the deep learning method. Firstly, the proband names were de-identified to ensure privacy. Subsequently, medical reports underwent various processes within the framework, including abbreviation expansion, translation correction, PHI de-identification according to the University of Chicago’s HIPAA privacy rules. The physician conducted a meticulous comparison between the results obtained from the pipeline and their own assessments. The evaluation involved assessing the quantity and accuracy of de-identification, translation, and summarization using HPO terms, while systematically documenting any errors encountered, specifying their severity, and providing explanations for their occurrence.

The primary endpoint of this study is to compare the number of PHI that are de-identify by an automated framework to the number that are de-identified by a physician.

The software validation for this study’s secondary endpoint includes evaluating the non-inferiority of the tool compared to a physician in terms of de-identification and symptom summarization in HPO format. For being sustainable in routine clinical practice, we targeted a minimal sensitivity of 90% and specificity of 75% of de-identification and a sensitivity of 75% and specificity of 90% of summarization. Errors in the de-identification process are categorized as major (*e.g*. including identifiable information) or moderate (*e.g*. indirectly identifying PHI), while errors in summarized information are categorized as major (*e.g.* missing the diagnosis) or moderate (*e.g*. incorrect summarized information). This three-levels assessing risk system is detailed in <u>Table S1</u>.

### DEEP LEARNING MEDICAL REPORT PROCESSING

Report was processed according to the following offline pipeline : a) Text was first expanded with abbreviations gathered previously, b) Expanded text was translated into English using the Marian translator, an open-source neural machine translation tool developed by OpenNMT ^11^ (https://marian-nmt.github.io), c) Translated and corrected text was de-identified from PHI using Microsoft’s Presidio (https://microsoft.github.io/presidio/), d) Translated and de-identified text is summarized in HPO format, including symptoms’ quantitative data, using a upgraded version of ClinPhen ^12^, and flagged “high-confidence” or “low-confidence” according the possibility of being associated to a relatives-concerned or a negatives sentences. An example of the deep learning method processing on a typical French medical report is presented in the Figure 1.

**Figure 1.**
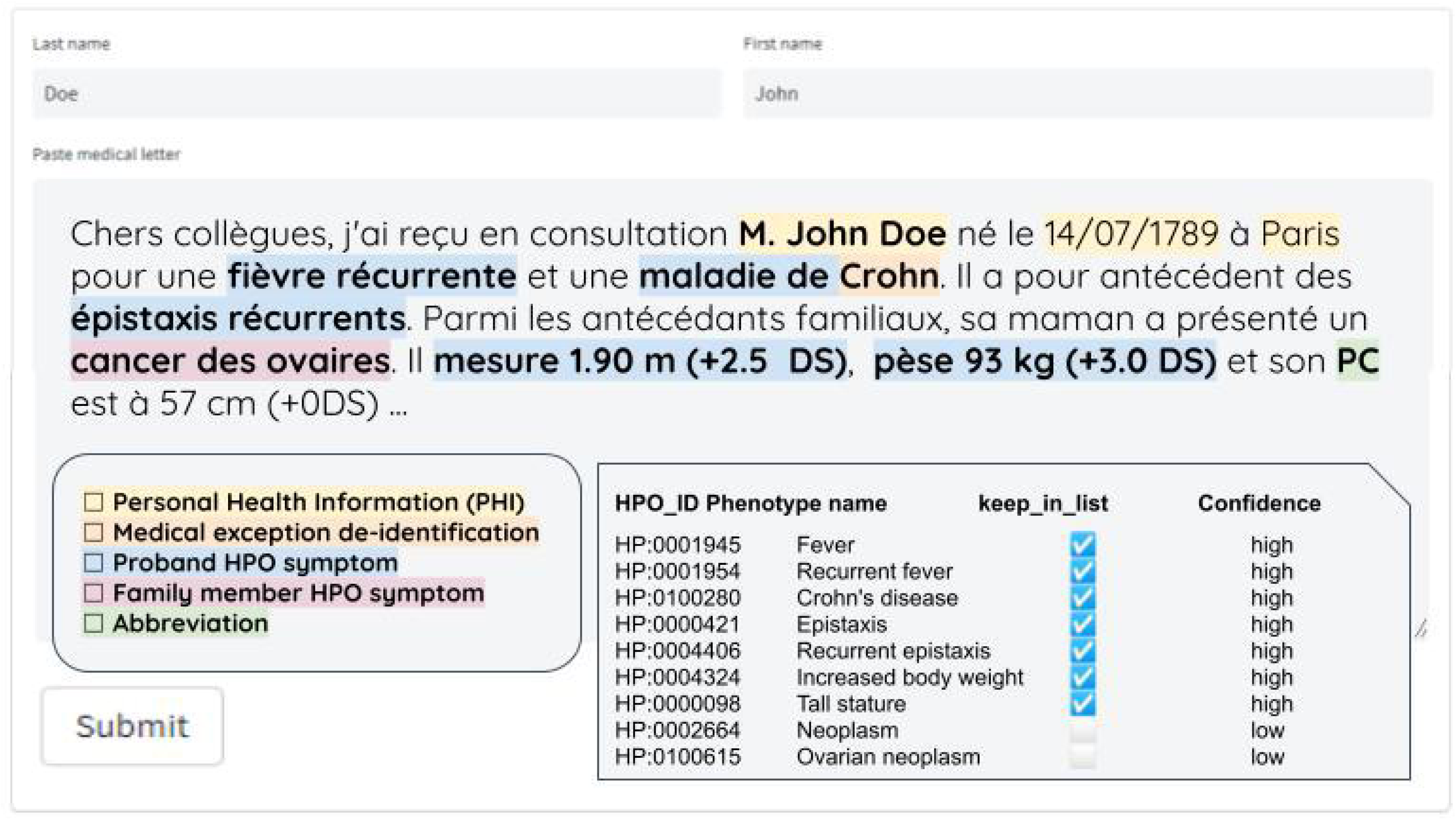
Deep learning framework example illustration. Example of a French medical report, encompassing personal health informations in yellow, medical proper name to exclude from de-identification in orange, proband Human Phenotype Ontology (HPO) symptom in blue, family member HPO symptom in pink and abbreviation in green, summarized into HPO terms with confidence level flags.

#### Hybrid processing with human correction of medical terms and databases

To avoid reading difficulties and losing clinical information, we implemented a hybrid process involving both human review and AI techniques <u>(Table S2)</u>, using French as a proof of concept. In order to identify and prevent de-identification errors, we compiled a comprehensive list of 2998 medical associated proper names, 4939 officinal drugs and 109804 genes symbols from various databases, including the OMIM phenotype title (March 2023 update, https://omim.org/), Human Phenotype Ontology terms (March 2023 release), ANSM lists of official medication in allopathy without homeopathy and herbal medicine (March 2023 update), and complete HGNC approved gene dataset (March 2023 update) to excluded them from de-identification. Given that method location detection was insufficient concerning French territories, we implemented a list composed of 34981 propers names of French cities, departments and regions coming from INSEE’ official geographic code (https://www.insee.fr/fr/information/6051727, January 2022 release) to force de-identification. Concerning abbreviation, we have listed 146 frequent French country-specific abbreviations, usually stand for pathologies, medical related structures, technologies and analysis, collected from Geneva University Hospitals’ glossary (http://abreviationsmedicales.ch/), as well as the collaborative Wikipedia page “Liste_d’abréviations en médecine” (https://fr.wikipedia.org/wiki/Liste_d%27abr%C3%A9viations_en_m%C3%A9decine), and the Pays de la Loire Regional Health Agency (ARS) abbreviation list (https://www.pays-de-la-loire.ars.sante.fr/system/files/2018-06/Aide%20-%20Acronymes.pdf), to write them out in full in French and translate them into English. Five among those cannot be expanded because of their ambivalent meaning (*e.g.* TCA, CMT, RCP). To ensure an accurate translation of the clinical information, a correction dictionary composed of 4646 terms was created from the French translation of Human Phenotype Ontology (accessible in PhenoTips website https://nexus.phenotips.org/nexus/content/repositories/releases/org/phenotips/vocabulary-hpo-translation-french/1.4-rc-4/, October 2018 release) and manually reviewed. 163 translations have been removed because of misleading translation without alternative, 54 have been replaced.

### STATISTICAL ANALYSIS

In this study, we have considered the missed PHI de-identification as False Negative (FN) and the excess of de-identification of non-PHI information as False Positive (FP). As well, for the summarization, we have considered the excess of HPO terms as False Positive and the missed HPO terms as False Negative. True Positive (TP) and True Negative (TN) were de-identification and summarization for which the assessing physician corroborates the tool statement. To assess this method, we use sensitivity and specificity thresholds as primary objectives. Specificity is a statistical measure that quantifies the proportion of true negatives in a diagnostic test, represented by the formula: Specificity = TN / (TN + FP).

Sensitivity is a statistical measure that quantifies the proportion of true positives in a diagnostic test, represented by the formula: Sensitivity = TP / (TP + FN).

### CODE AVAILABILITY

This deep learning method is open-source and can be installed for offline usage on a local machine by following instructions at https://github.com/kyauy/ClinFly or used directly online at https://huggingface.co/spaces/kyauy/ClinFly. This method manages French, German and Spanish language. A translated and de-identified report can be downloaded as a text file output. The list of symptoms can be downloaded in CSV or PhenoTips JSON format, compatible with most EHR entries especially in genome sequencing platforms.

## Results

### STUDY DATA

From March to April 2023, we have gathered 46 consultations reports and 4 hospitalization reports written between 2019 and 2023 by eight differents physicians, concerning 50 patients, whose median age in report was 6 years old coming from 12 differents indications of which neurodevelopmental disorders (54%), congenital disorders, fetal pathology and oncology. In median, a report contains 478 words, 3180 characters, 3 abbreviations, 15 PHI and 7 HPO terms. The distribution between the PHI and HPO terms is shown in Figure 2. The cohort characteristics are detailed in the Table 1.

**Figure 2.**
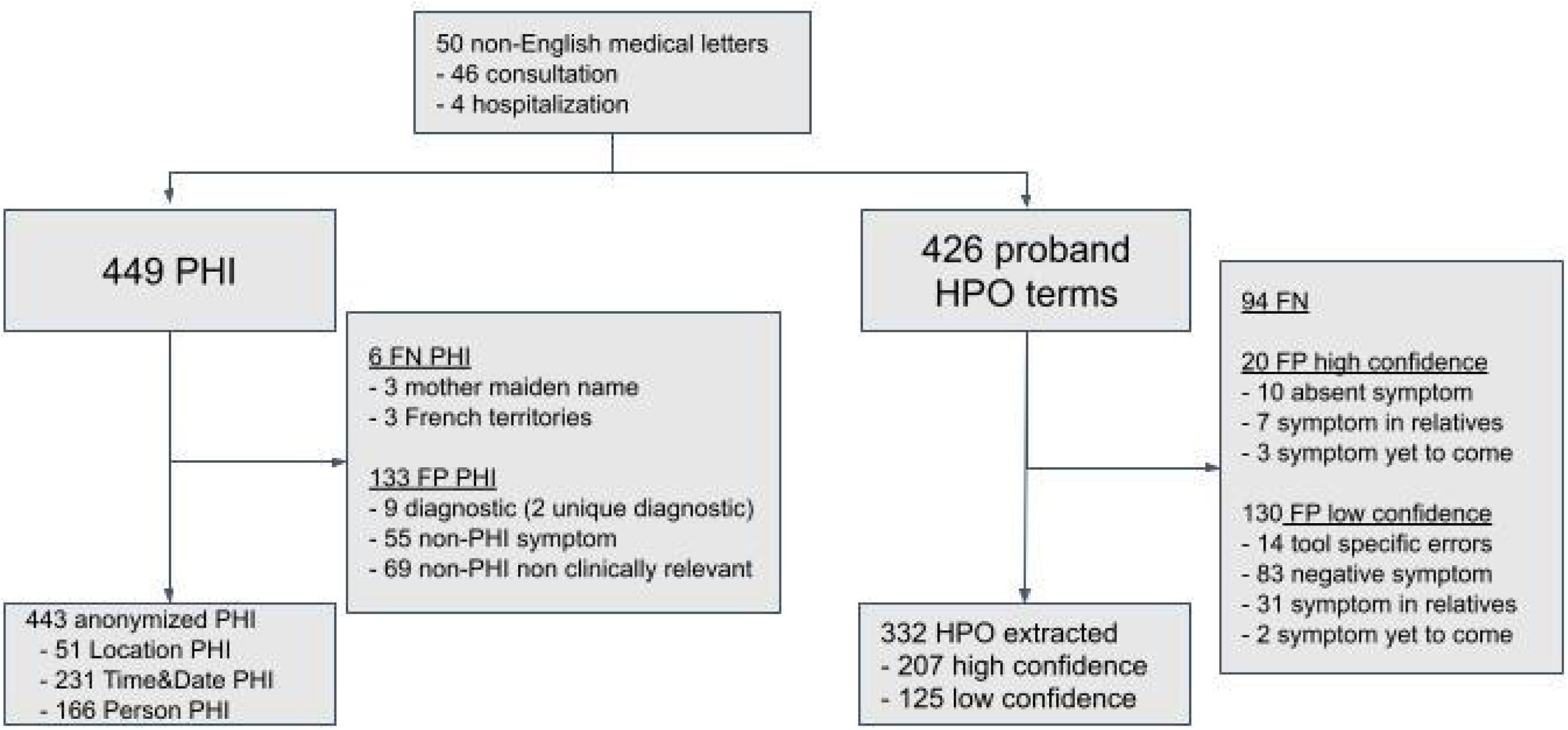
Overview of the non inferiority trial analysis. Protected Health Information de-identification and summarization in Human Phenotype Ontology (HPO) symptoms of non-English medical reports.

**Table 1.** Study cohort. Characteristics of the series medical reports.

| Cohort | n [min_max] |
| --- | --- |
| Number of physician | 8 |
| Number of consultation letter | 50 |
| Number of indication | 12 |
| Median age in letter | 6 [0-50] |
| Metrics per letter in median | n [min_max] |
| Word | 478 [229-1728] |
| Characters | 3180 [1571-11405] |
| Abbreviations | 3 [0-14] |
| PHI | 15 [5-70] |
| HPO terms | 7 [2-26] |

### STUDY ANALYSIS

#### Primary endpoint

Deep learning method de-identified 443 of the 449 existing PHI, achieving the primary objectives with a sensibility of 99%, which is superior to the 90% predefined minimal sensibility threshold. Concerning de-identification, 51 were due to the French territories dictionary, 231 were due to the date and time detection module and 166 were due to the person detection module of the software. For the six remaining errors, three of those were moderate errors, missing a location information (*i.e.* “Paris”, “Nîmes”) and the three others were major errors, missing the mother’ maiden name of the proband.

#### Secondary endpoints

##### Assessing medical report summary using HPO format

Deep learning method summarized 207 on 227 high confidence correct HPO terms, reaching a specificity of 92%, which is superior to the 90% predefined minimal specificity threshold of summarizing medical report using HPO format. The low confidence summarized informations allowed to save 125 HPO terms, on 255 proposed, leading to 29% of recall. Encompassing the high and low confidence summarized informations, deep learning method summarized 332 on the 426 HPO terms, which is equivalent to 6.6 HPO per report in average, and an overall sensibility of 78%, which is superior to the 75% predefined minimal sensibility threshold.

The 130 low confidence HPO terms left were objectively false because of a negative form (83/130), a family member sentence (31/130), a translation/summarized issue (130) or due to a mention of a symptom yet to come (2/130). Concerning the false high confidence HPO terms summarization, half of them were because of a translation or an summarization issue (major error), 7 of them were due to a family member sentence (moderate error) and 3 of them were due to the mention of symptoms yet to come, in description syndrome paragraph (not included as error).

##### Evaluation of de-identification processing

Deep learning method keeped 362 of 495 non-PHI informations, with an overall specificity of 78%, or a 87% specificity if encompassing only moderate and major errors, which is superior to the 75% threshold.

Thanks to the hybrid processing to keep pertinent clinical information, deep learning only de-identified 9 non-PHI diagnosis or indication in excess (*i.e.* “onychodysplasia”, “Iso Kikuchi”) coming from the same medical report, 55 non-PHI clinical informations (symptom), 69 non-PHI non-clinically relevant words (this, by, morning…), considered respectively as major, moderate and minor errors.

## Discussion

In this study, we conducted a non-inferiority trial to evaluate the feasability and the risk of a deep learning method for translation, de-identification and summarization of medical reports compliant to health data privacy, compared to a physician.

To assess deep learning methods, we developed an open-source software reaching 99% sensibility and 87% specificity of de-identification of PHI information. Assessing medical reports summarizing using HPO format reaching a 92% specificity, highlighting the high reliability of the symptoms summarization using this system. Overall, this deep learning method successfully summarized 332 of the 426 HPO terms which is equivalent to 6.6 HPO per report in average. This performance is comparable to the average number of symptoms manually filled out by physicians in PhenoDB and the MatchMaker Exchange initiative ^13^.

To our knowledge, there is no other existing multi-language framework allowing physicians to translate, de-identify and summarize medical reports. Moreover, concerning the HPO format summarization, performances are similar to pre-existing automated tools such as Doc2HPO ^14^. This deep learning method reliably delivers scalable results, validated through rigorous testing, making it suitable for handling large data volumes without compromising accuracy. To ensure anonymity and accuracy, the deep learning framework was specifically designed to maintain high sensitivity in PHI de-identification and high specificity in summarizing medical reports using HPO, prioritizing excess of de-identification of the medical report and generating fewer but accurate symptoms. These sensitivity and specificity thresholds were chosen because they allow the framework to be used in routine clinical practice, although human corrective action maybe necessary to verify the data generated especially to get the most comprehensive data. Despite extensive efforts, some PHI remained identifying. However, we believe that these remaining location-related PHI can be effectively addressed through software enhancements, further improving the efficiency of deep learning in de-identify medical reports. This method does process German and Spanish medical reports, however, no manual curation improvement has been developed for those languages yet. Our study data was mainly based on medical reports from clinical geneticists. Therefore this could explain why family member symptoms represent 35% of excess of summarized informations errors ; we consider that this risk is maximal in genetic medical reports due to the systematic mention of the family medical background.

In addition, proper names from classifications, surgical techniques and study names are not managed and usually undergo an excess of de-identification. We did not implement a module to recognize drug medications indications and summarize by the corresponding HPO terms. Another limitation of this method is the management of multiple patients and family members on a single medical report. Although we conducted preliminary evaluations and selected the tools based on our assessments, a more comprehensive benchmark of existing deep learning methods would have been beneficial.

Machine learning tools such as LLM and GPT could benefit for translation, de-identification and summarization when it will be compatible with medical data privacy restriction ^15^.

Overall, our results demonstrate the potential of AI methods in improving the efficiency and accuracy of PHI de-identification and summarizing medical reports in HPO format. AI could help to overcome manual entries of clinical data, facilitating the exploitation of the “genotype-first” approach in medical genomics to discover new disease-gene, expand clinical spectrum and retro-phenotype the patient by presume and confirm the diagnosis with a variant of interest.

Deep learning method reports non-inferior performance as physician to de-identify and summarize in HPO format non-English medical reports with reliable and scalable results. This study provides insights on how to exploit medical reports that allows physicians to share structured clinical data to facilitate precision medicine.

## Data Availability

All data produced in the present study are available upon reasonable request to the authors

https://huggingface.co/spaces/kyauy/ClinFly

## Supplementary material and methods

### Supplementary methods

#### Methods S1. Additional informations on medical reports processing using ClinFly

To ensure accuracy, we implemented specific measures such as strict separation of abbreviations with spaces during expansion.Furthermore, we replaced the term “associated” with “with” to avoid potential errors resulting from considering it as indicative of a disease name. Additionally, punctuation-related issues prompted us to replace abbreviations like “M.,” “Dr.,” and “Pr.” with appropriate alternatives.

To ensure the preservation of clinical information, we implemented specific exclusions for the de-identification of dates and times : this involved retaining temporal words (e.g., years, months, noon, hours) and slash-related information (e.g., blood pressure 140/80, Apgar score 7/7/8/10) that do not represent specific dates (e.g., 10/12/1994). Considering the low likelihood of license plate and number mentions in medical reports, we deactivated their detection to minimize the risk of misidentifying relevant information (e.g., genes, height). Furthermore, to maintain anonymity, the proband’s first and last names were pseudonymized as “CAS” and “INDEX,” respectively, ensuring clear distinction from other anonymous individuals.

ClinFly utilizes a clause-by-clause analysis approach, employing punctuation marks such as commas and periods to effectively manage false-positive summarization, particularly from negative sentences and information related to the patient’s family members. Our systematic summarization method focuses on the first sentence of the consultation letter, which commonly contains the primary reason for the patient’s visit, while also considering any relevant clinical details pertaining to their relatives. Notably, an upgraded version of ClinPhen was developed to address errors related to HPO_ID synonyms.

### Supplementary Tables

Table S1. Assessing risk system of using deep-learning de-identification and summarization of medical letters in three-levels of severity

Table S2. Curated resources for hybrid deep learning method describing the collection of human-reviewed dictionaries included in the process for managing translation and de-identification issues. The table highlights the content count, required sources, and provides an example of the dictionaries’ functionality.

