## Supplementary figures and images for "Assessing feasibility and risk to translate, de-identify and summarize medical reports using deep learning"

### Table S1

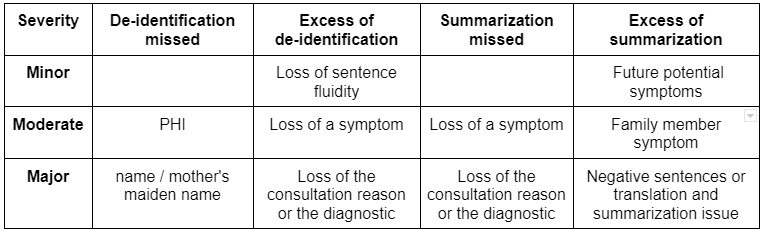

### Table S2

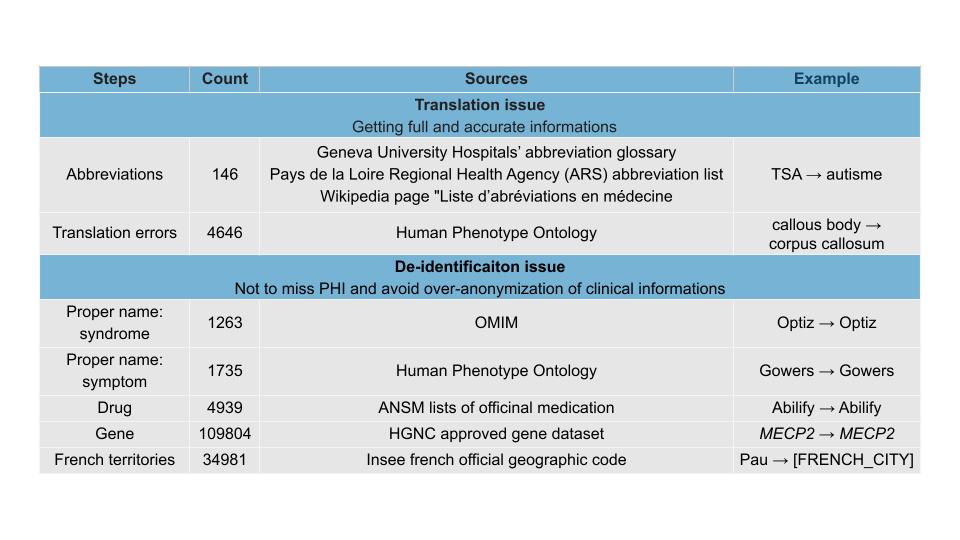
